# Pseudotemporal whole blood transcriptional profiling of COVID-19 patients stratified by clinical severity reveals differences in immune responses and possible role of monoamine oxidase B

**DOI:** 10.1101/2022.05.26.22274729

**Authors:** Claire Broderick, Irene Rivero Calle, Alberto Gómez Carballa, Jose Gómez-Rial, Ho Kwong Li, Ravi Mehta, Heather Jackson, Antonio Salas, Federico Martinón-Torres, Shiranee Sriskandan, Michael Levin, Myrsini Kaforou, the BioAID Consortium and GEN-COVID Study Group

## Abstract

Severe acute respiratory syndrome coronavirus 2 (SARS-CoV-2) infection is associated with highly variable clinical outcomes. Studying the temporal dynamics of host whole blood gene expression during SARS-CoV-2 infection can elucidate the biological processes that underlie these diverse clinical phenotypes. We employed a novel pseudotemporal approach using MaSigPro to model and compare the trajectories of whole blood transcriptomic responses in patients with mild, moderate and severe COVID-19 disease. We identified 5,267 genes significantly differentially expressed (SDE) over pseudotime and between severity groups and clustered these genes together based on pseudotemporal trends. Pathway analysis of these gene clusters revealed upregulation of multiple immune, coagulation, platelet and senescence pathways with increasing disease severity and downregulation of T cell, transcriptional and cellular metabolic pathways. The gene clusters exhibited differing pseudotemporal trends. Monoamine oxidase B was the top SDE gene, upregulated in severe>moderate>mild COVID-19 disease. This work provides new insights into the diversity of the host response to SARS-CoV-2 and disease severity and highlights the utility of pseudotemporal approaches in studying evolving immune responses to infectious diseases.

## 1 Introduction

Outcomes from severe acute respiratory syndrome coronavirus 2 (SARS-CoV-2) infection vary greatly from asymptomatic infection to severe, life-threatening multi-system disease. An exaggerated hyperinflammatory immune response with neutrophil and macrophage activation and “cytokine storm” is associated with severe clinical phenotypes [1-6], as well as impaired type 1 interferon signaling [7, 8] and T cell dysfunction [9-12], suggesting that severe disease is the outcome of a complex interplay between the various components of the immune response and SARS-CoV-2.

Studying the dynamics of the immune response to SARS-CoV-2 infection in differing severity phenotypes can provide crucial insights into the pathogenesis of severe COVID-19 disease and inform development of host-directed therapies and diagnostics. Time-course studies of anti-SARS-CoV-2 immune responses have thus far focused on proteomic and cellular approaches [13-19]. Longitudinal transcriptomic studies, which can provide valuable insights into the temporal patterns of immunological responses to infectious diseases [20] have been limited to date, and have focused on biomarker discovery [21] and comparisons of acute disease with convalescence and recovery [16, 22]. Longitudinal studies require the same patients to contribute sequential samples and can be logistically challenging. Pseudotemporal approaches, where each patient may contribute just one sample to the analysis, offer an alternative.

In this study we employed a novel pseudotemporal technique for exploring the trajectory of transcriptomic responses in COVID-19. A pseudotemporal model was constructed from patients’ single timepoint samples, and the trajectory of gene expression over pseudotime was modelled for mild, moderate and severe COVID-19 disease. Pseudotemporal differential analysis identified genes that were significantly differentially expressed (SDE) over pseudotime and between the different severity groups and pathway analysis was undertaken, aiming to elicit the temporal patterns of biological mechanisms underlying the different severity phenotypes.

## 2 Materials and Methods

### 2.1 Clinical cohort

Whole blood RNA-Sequencing (RNA-Seq) datasets arising from adults (age ≥18 years) presenting with SARS-CoV-2 infection in March to May 2020, were employed from Gene Expression Omnibus (adult COVID-19 and healthy controls from United Kingdom [UK]; adult COVID-19 and inflammatory bowel disease from Spain). The corresponding clinical metadata were reviewed, and clinical phenotyping was undertaken. Samples were excluded if there was known bacterial or fungal coinfection, haematological disorders or pre-existing immunosuppression including Human Immunodeficiency Virus (HIV), as these conditions were thought likely to impact the transcriptome.

All patients had been scored on the World Health Organization 8-point ordinal scale for assessing COVID-19 disease[23] Severity stratification was based on these scores, both for clinical status at the time of research blood sampling (“Severity Sample”) and for overall greatest severity during the COVID-19 episode (“Worst Severity”). “Mild” was defined as those who scored 1-2, were ambulatory and did not require hospitalisation for COVID-19; “Moderate” was those scored as WHO 3-4, requiring hospitalisation with or without standard oxygen therapy; “Severe” was defined as those scoring 5-8, who received at least one of high flow oxygen (>16 liters/ minute), non-invasive ventilation, mechanical ventilation, inotropes, haemofiltration, and/ or who died. Given the complexity of the planned analysis, it was important to minimise potential confounders. Thus, samples were excluded if “Severity Sample” differed from “Worst Severity”, or if patients had received immunomodulating treatment for COVID-19 prior to research blood sampling, including steroids, tocilizumab and interferon. “Symptom duration” was calculated as the number of days between symptom-onset and research blood sampling and each sample was then assigned a Pseudotime, based on symptom duration.

### 2.2 Statistical analysis

All statistical analyses were performed using the statistical software R (R version 4.0.4) [24]. The two RNA-Seq datasets were merged, keeping only those genes present in both datasets. Lowly expressed genes (total counts <10) and ribosomal genes were removed. Batch effect adjustment was performed using ComBat-seq (sva package version 3.38.0) [25]. Normalised counts were calculated for each gene with DESeq2 (V1.30.0) [26], using the default parameters. Normalised genes with fewer than three samples with a normalised read count of at least 20 were considered lowly expressed and were removed, leaving a total of 20,986 genes. Principle component analysis (PCA) was performed on the normalised counts for quality control and to confirm the removal of the batch effect.

Pseudotemporal differential expression analysis, to identify genes significantly differentially expressed (SDE) over pseudotime and between severity groups, was performed using MaSigPro [27], with “Mild” as the comparator group, two degrees of freedom to capture linear and quadratic trends and binomial distribution specified. MaSigPro follows a two-step regression strategy to find genes with significant temporal expression changes and significant differences between groups. Coefficients obtained in the second regression model were then used to cluster together significant genes with similar expression patterns. Calculation of adjusted p-values was performed using the Benjamini-Hochberg (BH) procedure [28]. Genes with an adjusted p-value < 0.05 were considered SDE.

To complement and validate the MaSigPro analysis, time-course differential expression analysis was performed in DESeq2, using the likelihood ratio test. SDE genes (BH-adjusted p-value < 0.05) identified were identified for the factors Pseudotime, Severity and Severity: Pseudotime.

Pathway analysis of SDE genes, in which genes are mapped to biological processes and pathways, was undertaken in gProfiler (https://biit.cs.ut.ee/gprofiler/gost). Reactome reference database was used and significantly enriched terms defined as those with adjusted p-value <0.05.

## 3 Results

### 3.1 Description of clinical cohort

Whole blood RNA sequencing data were available for 99 COVID-19 patients and 10 healthy controls (HC) from the UK, and 58 COVID-19 and 10 inflammatory bowel disease (IBD) patients from Spain, with each patient contributing one sample. Of these, 29 COVID-19 samples (22 UK, 7 Spain) were excluded due to presence of coinfection, haematological disorder or pre-existing immunosuppression. The remaining 128 COVID-19, 10 HC and 10 IBD samples were taken forwards for batch correction and normalisation. The HC and IBD samples, included to aid batch correction and normalisation, were treated as independent groups and were removed after these steps. A further 27 COVID-19 samples were excluded from the analysis due to prior treatment with immunomodulators (steroids, tocilizumab, interferon) for COVID-19 (n=11), Sample Severity category differing from Worst Severity category (n=15) or both (n=1). A further two samples from individuals who remained asymptomatic were excluded as they could not be assigned to a Pseudotime, as well as one sample which was an outlier with respect to symptom duration (42 days, 16 days longer than the next longest symptom duration) (Figure 1).

**Figure 1.**
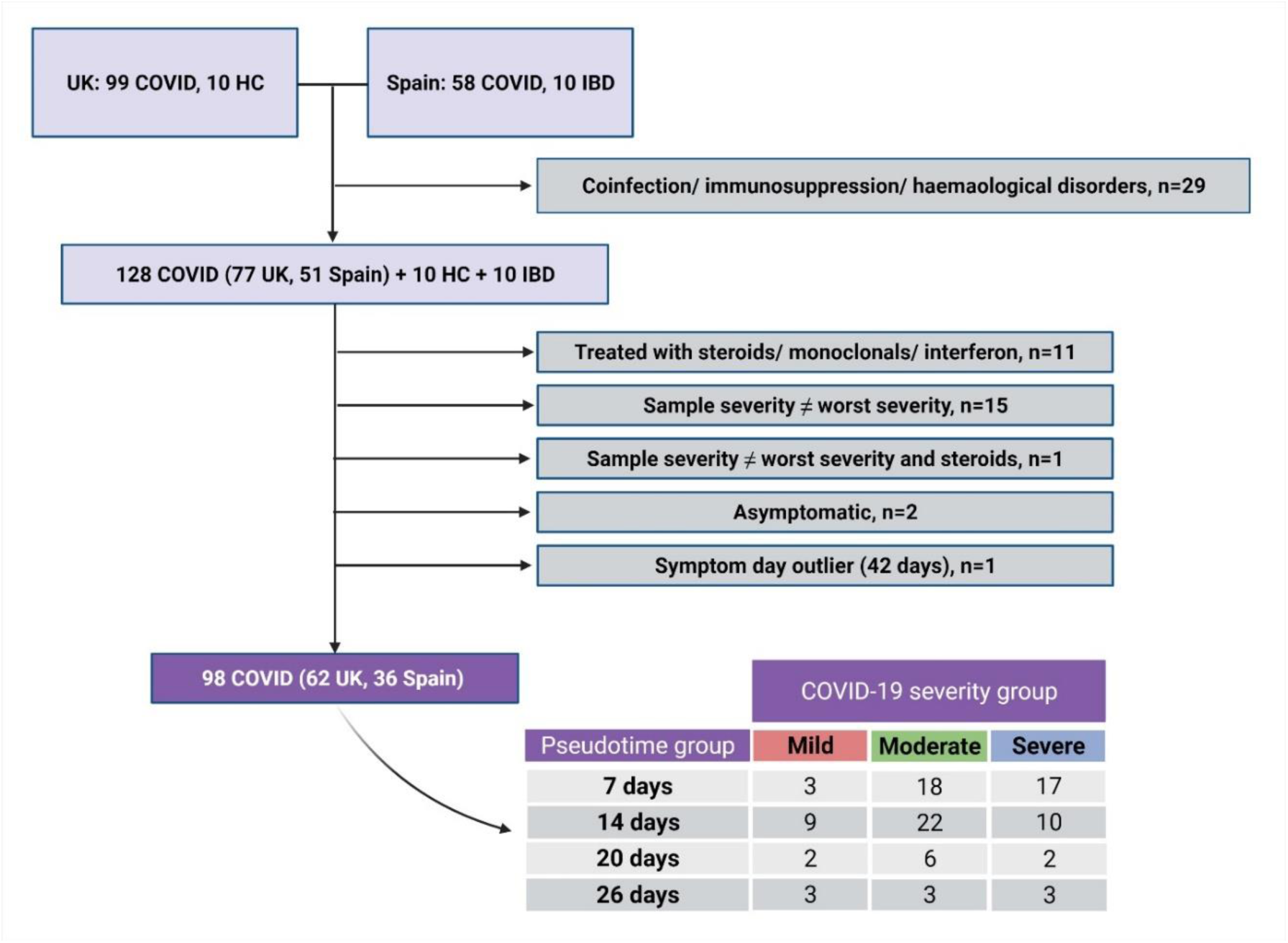
Summary of patient samples, inclusions and exclusions. Number of patients in each COVID-19 severity and pseudotemporal group is shown. Created with BioRender.com.

In total, 98 samples (62 UK, 36 Spain) were taken forwards to the pseudotemporal analysis, with 17 categorised as Mild (median age 37 years, 82% female), 49 Moderate (median age 61 years, 47% female) and 32 Severe (median age 68 years, 28% female). Each sample was also assigned a Pseudotime category based on symptom duration (number of days between symptom onset and research blood sampling): 1-7 days (d) symptom duration = 7 d pseudotime; 8-14 d symptom duration =14 d pseudotime; 15-20 d symptom duration =20 d pseudotime; 21-26 d symptom duration =26 d pseudotime (Figure 1, Supplementary table 1). These pseudotime values were inputted as the MaSigPro variable “Time”.

### 3.2 Pseudotemporal analysis of differential gene expression between COVID-19 severity groups

After batch correction and normalisation, principle component analysis confirmed that batch effects had been removed (Supplementary figure 1). The pseudotemporal differential expression analysis in MaSigPro identified 5,267 genes SDE between the severity groups and over pseudotime. To focus on those SDE genes which best fitted the model, an R^2^ threshold >0.2 was applied, giving 2,417 SDE genes, of which 2,405 were SDE between Severe *vs* Mild and 1,900 were SDE between Moderate *vs* Mild (Supplementary file 1). The top gene in the Severe *vs* Mild and Moderate *vs* Mild comparisons was MAOB (p=1.2×10^−18^, expression plotted in Figure 2A), followed by IGHV4-28, EGF, GAS2L1 and TRAV6. Coefficients obtained in these comparisons were used to cluster significant genes with similar pseudotemporal expression patterns, using hierarchical clustering. Four gene clusters were revealed with expression values highest in Severe > Moderate > Mild and two clusters with expression values highest in Mild > Moderate> Severe (Figure 2B and Supplementary file 2). Pseudotemporal patterns differed between clusters, for example in *clusters 1 and 4*, gene expression in Moderate and Severe were similar at early timepoints, before diverging after 14 days. By contrast, in *clusters 3 and 5*, gene expression appeared higher in Severe *vs* Moderate throughout pseudotime

**Figure 2.**
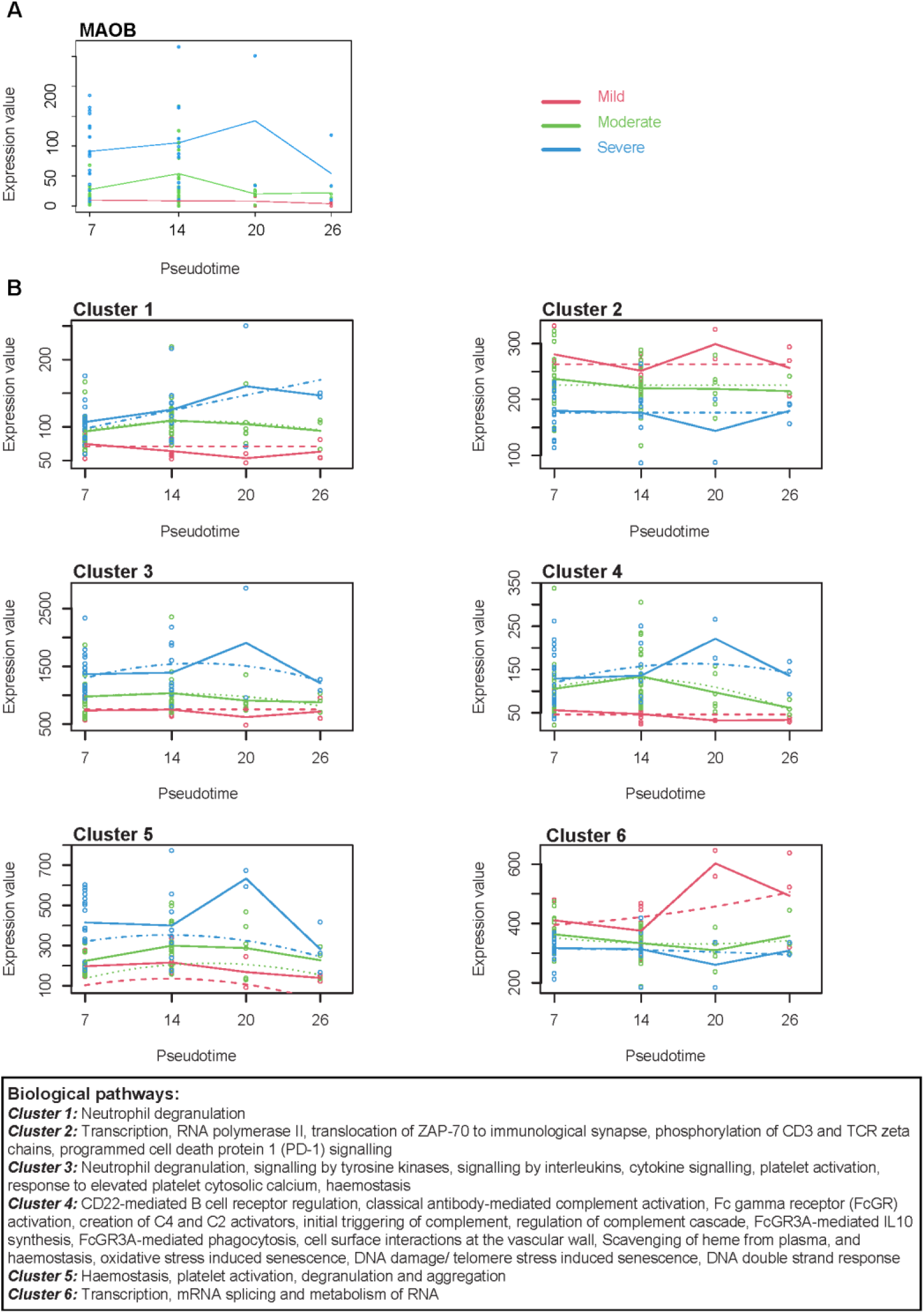
Pseudotemporal expression of monoamine oxidase B and gene clusters. In total, 5,267 genes were significantly differentially expressed (SDE) over pseudotime and between the severity groups (BH corrected p value < 0.05). (A) The top SDE gene was monoamine oxidase B (MAOB). (B) The coefficients obtained were used to group together SDE genes with R^2^>0.2 into 6 clusters with similar pseudotemporal expression patterns. Plots of gene expression over pseudotime for the Mild (red), Moderate (green) and Severe (blue) groups are shown. Solid lines connect the median expression to show the trends for each group, and the dashed lines show the regression curves fitted to the data.

Pathway analysis of genes in the four clusters with highest expression in Severe samples (1, 3, 4, 5) found significant enrichment for immunological and coagulation pathways. *Cluster 1* was enriched for neutrophil degranulation. *Cluster 3* was enriched for neutrophil degranulation, signalling by tyrosine kinases, signalling by interleukins, cytokine signalling, platelet activation, response to elevated platelet cytosolic calcium and haemostasis. *Cluster 4* was enriched for multiple immunological and haemostasis pathways included CD22-mediated B cell receptor regulation, classical antibody-mediated complement activation, Fc gamma receptor (FcGR) activation, Creation of C4 and C2 activators, Initial triggering of complement, regulation of complement cascade, FcGR3A-mediated interleukin-10 synthesis, FcGR3A-mediated phagocytosis, cell surface interactions at the vascular wall, Scavenging of heme from plasma, and haemostasis. Senescence, Cell cycle and DNA repair pathways were also significantly enriched in *Cluster 4*, including Oxidative Stress Induced Senescence, DNA Damage/Telomere Stress Induced Senescence, DNA double strand break response, cleavage of damaged purines and base excision repair. *Cluster 5* was enriched for coagulation pathways including haemostasis, platelet activation, degranulation and aggregation and formation of fibrin clot (clotting cascade). (Supplementary file 3 for pathways lists for each gene cluster).

Two clusters (2, 6) had expression lowest in the Severe samples. Cluster 2 was enriched for transcription, RNA polymerase II, translocation of ZAP-70 to immunological synapse, phosphorylation of CD3 and TCR zeta chains and programmed cell death protein 1 (PD-1) signalling. Cluster 6 was enriched for transcription, mRNA splicing and metabolism of RNA. (Supplementary file 3).

### 3.3 Time series analysis in DESeq2

As an alternative method for evaluating genes differentially expressed over pseudotime and severity, a time series analysis was conducted in DESeq2. In total, 6,042 were SDE with Severity and 275 SDE with Pseudotime. There were 61 genes SDE with the interaction Severity: Pseudotime, all 61 of which overlapped with both the Severity and Pseudotime SDE genes (Supplementary file 4, Figure 3). Of the 5,136 SDE genes identified in the MaSigPro analysis, 4,612 were identified as SDE in the DESeq2 analysis (Figure 3), confirming the MaSigPro results.

**Figure 3.**
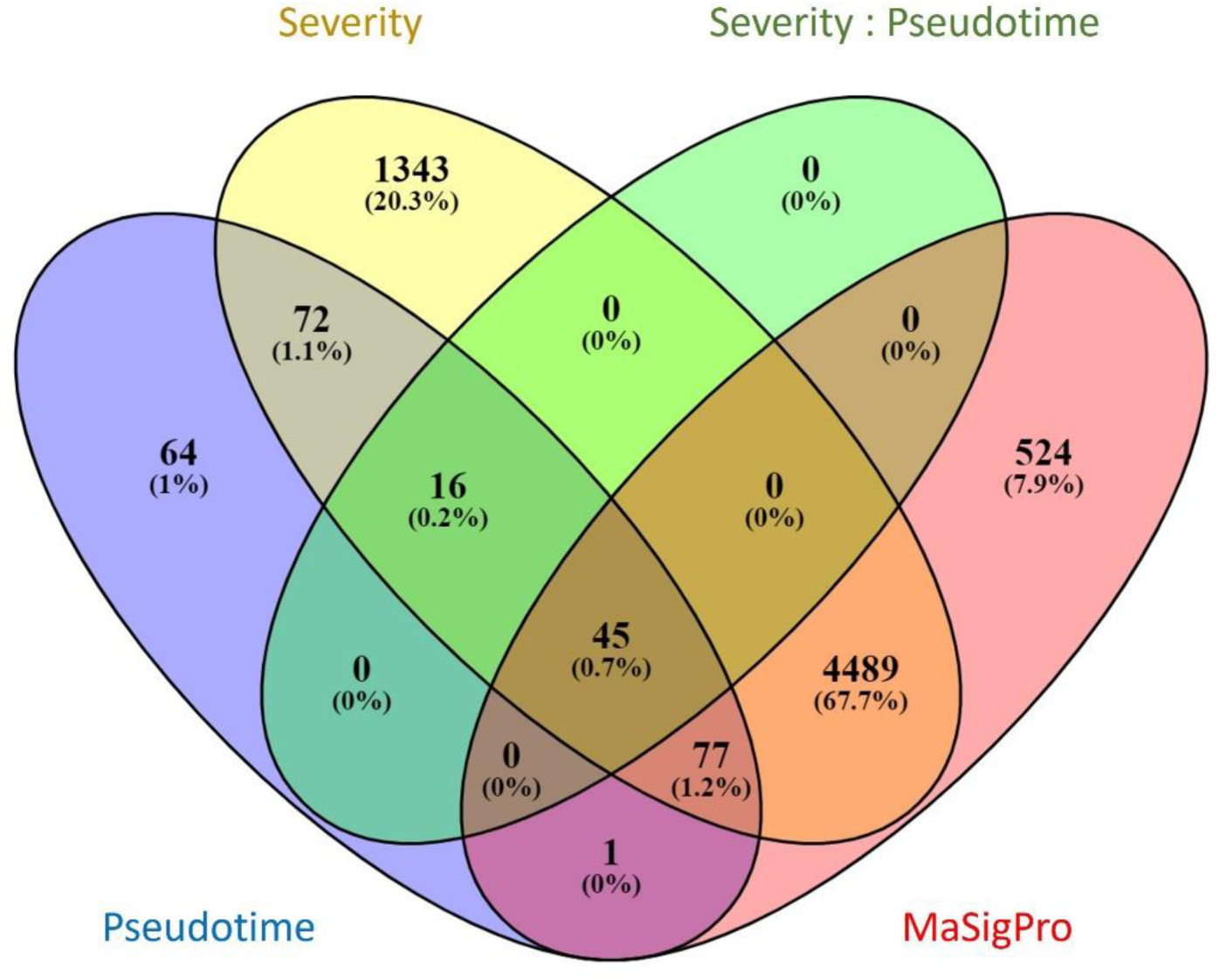
Venn diagram showing overlap of significantly differentially expressed genes from the DESeq2 (with Severity, Pseudotime, Severity: Pseudotime) and MaSigPro analyses.

## 4 Discussion

We have employed a novel pseudotemporal approach to explore trends in whole blood gene expression in patients with varying COVID-19 severity phenotypes. We have identified genes significantly differentially expressed over pseudotime and between severity groups, clustered these genes together based on pseudotemporal trends and undertaken pathways analysis of these gene clusters. Our analysis revealed that increasing severity was associated with upregulation of multiple immunological, coagulation and platelet pathways and downregulation of RNA metabolism, transcription and cellular metabolic pathways. We observed differing pseudotemporal trends: for example, there was divergent expression of genes enriched for coagulation and platelet pathways between severe and moderate groups from the earliest pseudo-timepoints, whereas expression of genes enriched for humoral immunological pathways were similar between these groups at early pseudo-timepoints before diverging after 14 days.

We observed upregulation of neutrophil activation pathways, highest in severe >moderate > mild clinical phenotypes (*Clusters 1 and 3*). Peripheral neutrophilia is a feature of severe COVID-19 [29] and bulk [1, 30] and single-cell [31] RNAseq studies have associated neutrophil pathways with COVID-19 severity. Neutrophil extracellular traps (NETs), networks of chromatin and bactericidal proteins released by neutrophils in response to pathogens, have been implicated in the pathogenesis of severe COVID-19 disease [32, 33]. In this study, differences in the expression of genes enriched in neutrophil pathways were observed throughout pseudotime. This suggests that neutrophils play an early role in severe disease and also have a continuing role in those with ongoing severe disease. This is consistent with longitudinal whole blood immune profiling of COVID-19 patients [15].

Coagulation pathways and platelet activation and degranulation pathways were upregulated in severe >moderate > mild clinical phenotypes (*Cluster 5*). COVID-19 disease is associated with a hypercoagulable state and abnormal coagulation parameters are associated with poor outcome [34]. Arterial and venous thromboembolism are important life-threatening complications of COVID-19 disease [35]. There is growing evidence for the role of platelets in COVID-19-associated hypercoagulability and acute respiratory distress syndrome (ARDS). Blood from hospitalised COVID-19 patients demonstrates increased platelet activation, aggregation and degranulation compared to healthy controls, with greater platelet hyperreactivity observed in those requiring intensive care [36-39]. SARS-CoV2 infection alters the platelet transcriptome, with gene expression changes in pathways associated with protein ubiquitination, antigen presentation, and mitochondrial dysfunction. Increased circulating platelet-neutrophil aggregates observed in hospitalised COVID-19 patients, particularly those in ICU, may contribute to the acute lung damage and ARDS [38]. In our study, there was differential expression of *Cluster 5* between the severity groups throughout pseudotime suggesting that disturbances in clotting and platelet pathways appear early and persist with ongoing severe disease. Interestingly, just one of the study participants (Moderate severity) had documented thrombosis.

We observed enrichment for B cell receptor pathways, complement activation and FcGR signalling pathways (*Cluster 4*) with greater expression in severe >moderate > mild clinical phenotypes. Multiple immunoglobulin genes contributed to this gene cluster, 41/48 of which are immunoglobulin variable genes. IGHV4-28 was also the second most significant gene in both the Severe *vs* Mild and Moderate *vs* Mild comparisons. These findings are consistent with the observation that anti-SARS-CoV-2 antibody levels positively correlate with increasing disease severity [40, 41]. B cell receptor sequencing has demonstrated clonal expansions in severe COVID-19 patients compared with non-severe patients and healthy donors. Overrepresentation of IGHV3-7, which is one of the *Cluster 4* SDE genes in our analysis, and IGKV3-15 have been observed in severe patients[42]. These observations raise the possibility that specific immunoglobulins could be involved in promoting severe disease, for example through Fc-gamma receptor activation of neutrophils and macrophages or complement pathways.

*Cluster 4* was also enriched for senescence pathways. Viral infection of cells can trigger cellular senescence (Virus-induced senescence)[43, 44]. Senescent cells secrete pro-inflammatory cytokines, extracellular matrix factors and pro-thrombotic mediators[45, 46] as part of a “senescence-associated secretory phenotype”. Recently published in vivo and in vitro work has suggested SARS-CoV-2 VIS as a possible trigger of COVID-19 cytokine storm and organ damage, and a potential therapeutic target. Supernatants from VIS cells induced formation of NETs, activated platelets and coagulation pathways. VIS was also associated with macrophage activation and complement lysis. Senolytics reduced inflammation and lung disease in SARS-CoV-2 infected mice[47].

Two gene clusters demonstrated reduced expression in severe < moderate< mild disease (*Clusters 2 and 6*), and on pathway analysis were found to be enriched for transcription and cellular metabolic pathways. Infection with SARS-CoV-2 virus suppresses host cell transcription and disrupts protein trafficking [48, 49], suppressing host innate anti-viral responses. We also observed reduced expression of pathways associated with T cell activation and signaling in the severe COVID-19 patients in *Cluster 2* (translocation of ZAP-70 to immunological synapse, phosphorylation of CD3 and TCR zeta chains and PD-1 signalling). T cell depletion and functional exhaustion are features of COVID-19, particularly in severe disease and are associated with PD-1 expression [11]. The PDCD1 gene which encodes PD-1 was not SDE in our MaSigPro analysis. ZAP-70 is a critical kinase in TCR signalling and T-cell activation[50]. Reduced phosphorylation of key proteins in the CD45/TCR signalling/ activation pathway, including ZAP-70, has been observed in the peripheral blood mononuclear cells (PBMCs) of hospitalised COVID-19 patients compared to healthy controls, with phosphorylation restored through the addition of C24D peptide [51]. Expression of PD-1 can be induced in multiple immunological cells, particularly T cells. In acute infection it can serve as a marker of T cell activation, whereas in chronic infection, continued PD-1 expression is associated with dysfunctional, exhausted T cells. PD-1 is also necessary for the recovery of exhausted T cells. One of its inhibitory effects on T cell function is via reduced phosphorylation of ZAP-70/ CD3-zeta[52].

The finding of Monoamine oxidase B (MAOB) as the top SDE gene in the pseudotemporal differential analysis is interesting, with expression higher in severe> moderate> mild throughout pseudotime (Figure 2A). The gene encodes the enzyme monoamine oxidase B (MAO-B), which is one of two monoamine oxidase enzymes found in the outer mitochondrial membranes, which catalyze the oxidative deamination of monoamine neurotransmitters. MAO-B is found in the central nervous system, where increased activity is associated with neurodegeneration and Parkinson’s disease, and also found in platelets. Aging, diabetes and obesity have been associated with higher levels of monoamine oxidases [53-55], and they have been implicated in oxidative stress and endothelial dysfunction[56-58]. A recent study found an association between MAO-B activity and post-operative delirium. In the same study, computational analyses found a high degree of structural similarity between MAO-B and the SARS-CoV-2 spike protein binding region of ACE2 and suggested the potential for SARS-CoV-2 binding to interfere with MAO-B activity [59]. A further computational study, published as a pre-print, reported that the SARS-CoV-2 spike protein has similar affinity for the MAO enzymes as is it does for the ACE2 receptor and thus SARS-CoV-2 infection could interfere with MAO activity [60]. The authors suggest a potential role of MAO-B in the neurological and platelet-based complications of SARS-CoV-2 infection.

Longitudinal studies, which provide valuable information about evolving immunological responses to infectious diseases, are constrained by the need to collect multiple sequential samples from patients, which can be logistically challenging. Our study demonstrates that pseudotemporal approaches can be an informative alternative when sequential samples are unavailable.

To the best of our knowledge, this is the first reported analysis of pseudotemporal transcriptomic trends in SARS-CoV-2 infection with comparisons of severity phenotypes, however it has some limitations. There were differences in age and sex between the severity groups, with age and male sex increasing with severity, which is in keeping with the epidemiology of COVID-19 disease [61, 62]. Therefore it is possible that some of the SDE genes we have identified are driven by age or sex, rather than COVID-19 severity. However, given COVID-19 severity, age and sex are so closely intertwined, adjusting for these two variables could mask key drivers of severity, and thus our unadjusted analysis may be a more sensitive approach. This study combines data from UK and Spanish cohorts. Both cohorts were recruited and sampled during the first wave in early 2020, but there may have been differences between the two countries, for example in government advice for staying at home and clinical management. The complexity of this analysis required us to minimise potential interference of the transcriptome by variables such as COVID-19 treatments and coinfections. Therefore a strict set of pre-determined exclusion criteria were employed that resulted in just two thirds of the samples being included in the analysis. Thus the sample size in some of the later severity-pseudotime groups was modest. We only included samples for which “Sample Severity” and “Worst Severity” classification were the same. Therefore, the results of this analysis cannot inform predictions of severity or prognosis.

### 4.1 Conclusion

We have modelled and compared the pseudotemporal trajectories of whole blood transcriptomic responses in patients with mild, moderate and severe COVID-19 disease. We have observed that increasing disease severity is associated with upregulation of multiple immune pathways, including neutrophil, complement and immunoglobulin-mediated responses, as well as platelet activation and coagulation pathways, with varying pseudotemporal patterns identified over the clinical course. A potential role of senescence is also suggested. This work highlights the complex interplay between immunological, thrombotic and cellular factors that underlie the clinical spectrum of COVID-19 disease. It demonstrates the utility of pseudotemporal approaches in studying host-pathogen responses.

## Data Availability

Data produced in the present study are available upon reasonable request to the authors.

## Author Contributions

Conceptualisation: CB, MK; Data curation: CB, IRC, HK-L, RM, HJ, AG-C; Formal analysis: CB, HJ; funding acquisition: CB, IRC, AGC, HK-L, AS, FM-T, SS, ML, MK; Investigation: IRC, AG-C, HK-L, RM; Methodology: CB, HJ, MK; Project administration: CB, MK; Resources: IRC, AG-C, FM-T, SS, ML, MK; Software: CB, HJ; Supervision: SS, FMT, ML, MK; Validation: CB; Visualisation: CB; Writing—original draft: CB, MK; Writing—review and editing: ALL.

All authors have read and agreed to the published version of the manuscript.

## Acknowledgments

Many thanks to the patients and their families who took part in the studies from which this data originated.

## References

[1.] Aschenbrenner AC, Mouktaroudi M, Krämer B, Oestreich M, Antonakos N, Nuesch-Germano M, et al. Disease severity-specific neutrophil signatures in blood transcriptomes stratify COVID-19 patients. Genome Medicine. 2021;13(1):7.

[2.] Del Valle DM, Kim-Schulze S, Huang H-H, Beckmann ND, Nirenberg S, Wang B, et al. An inflammatory cytokine signature predicts COVID-19 severity and survival. Nature Medicine. 2020;26(10):1636–43.

[3.] Su Y, Chen D, Yuan D, Lausted C, Choi J, Dai CL, et al. Multi-Omics Resolves a Sharp Disease-State Shift between Mild and Moderate COVID-19. Cell. 2020;183(6):1479–95.e20.

[4.] Chen G, Wu D, Guo W, Cao Y, Huang D, Wang H, et al. Clinical and immunological features of severe and moderate coronavirus disease 2019. J Clin Invest. 2020;130(5):2620–9.

[5.] Giamarellos-Bourboulis EJ, Netea MG, Rovina N, Akinosoglou K, Antoniadou A, Antonakos N, et al. Complex Immune Dysregulation in COVID-19 Patients with Severe Respiratory Failure. Cell host & microbe. 2020;27(6):992–1000.e3.

[6.] Akgun E, Tuzuner MB, Sahin B, Kilercik M, Kulah C, Cakiroglu HN, et al. Proteins associated with neutrophil degranulation are upregulated in nasopharyngeal swabs from SARS-CoV-2 patients. PloS one. 2020;15(10):e0240012.

[7.] Zhang Q, Bastard P, Liu Z, Le Pen J, Moncada-Velez M, Chen J, et al. Inborn errors of type I IFN immunity in patients with life-threatening COVID-19. Science. 2020;370(6515).

[8.] Bastard P, Rosen LB, Zhang Q, Michailidis E, Hoffmann HH, Zhang Y, et al. Autoantibodies against type I IFNs in patients with life-threatening COVID-19. Science. 2020;370(6515).

[9.] Zheng M, Gao Y, Wang G, Song G, Liu S, Sun D, et al. Functional exhaustion of antiviral lymphocytes in COVID-19 patients. Cellular & Molecular Immunology. 2020;17(5):533–5.

[10.] Chen G, Wu D, Guo W, Cao Y, Huang D, Wang H, et al. Clinical and immunological features of severe and moderate coronavirus disease 2019. The Journal of Clinical Investigation. 2020;130(5):2620–9.

[11.] Diao B, Wang C, Tan Y, Chen X, Liu Y, Ning L, et al. Reduction and Functional Exhaustion of T Cells in Patients With Coronavirus Disease 2019 (COVID-19). Frontiers in immunology. 2020;11:827.

[12.] Zheng H-Y, Zhang M, Yang C-X, Zhang N, Wang X-C, Yang X-P, et al. Elevated exhaustion levels and reduced functional diversity of T cells in peripheral blood may predict severe progression in COVID-19 patients. Cellular & Molecular Immunology. 2020;17(5):541–3.

[13.] Lucas C, Wong P, Klein J, Castro TBR, Silva J, Sundaram M, et al. Longitudinal analyses reveal immunological misfiring in severe COVID-19. Nature. 2020;584(7821):463–9.

[14.] Seow J, Graham C, Merrick B, Acors S, Pickering S, Steel KJA, et al. Longitudinal observation and decline of neutralizing antibody responses in the three months following SARS-CoV-2 infection in humans. Nature Microbiology. 2020;5(12):1598–607.

[15.] Mann ER, Menon M, Knight SB, Konkel JE, Jagger C, Shaw TN, et al. Longitudinal immune profiling reveals key myeloid signatures associated with COVID-19. Science Immunology. 2020;5(51):eabd6197.

[16.] Mathew D, Giles JR, Baxter AE, Oldridge DA, Greenplate AR, Wu JE, et al. Deep immune profiling of COVID-19 patients reveals distinct immunotypes with therapeutic implications. Science. 2020;369(6508):eabc8511.

[17.] Xu Z-S, Shu T, Kang L, Wu D, Zhou X, Liao B-W, et al. Temporal profiling of plasma cytokines, chemokines and growth factors from mild, severe and fatal COVID-19 patients. Signal Transduction and Targeted Therapy. 2020;5(1):100.

[18.] Liu J, Li S, Liu J, Liang B, Wang X, Wang H, et al. Longitudinal characteristics of lymphocyte responses and cytokine profiles in the peripheral blood of SARS-CoV-2 infected patients. EBioMedicine. 2020;55:102763.

[19.] Notarbartolo S, Ranzani V, Bandera A, Gruarin P, Bevilacqua V, Putignano AR, et al. Integrated longitudinal immunophenotypic, transcriptional and repertoire analyses delineate immune responses in COVID-19 patients. Sci Immunol. 2021;6(62).

[20.] Dunning J, Blankley S, Hoang LT, Cox M, Graham CM, James PL, et al. Progression of whole-blood transcriptional signatures from interferon-induced to neutrophil-associated patterns in severe influenza. Nature Immunology. 2018;19(6):625–35.

[21.] Gupta RK, Rosenheim J, Bell LC, Chandran A, Guerra-Assuncao JA, Pollara G, et al. Blood transcriptional biomarkers of acute viral infection for detection of pre-symptomatic SARS-CoV-2 infection: a nested, case-control diagnostic accuracy study. The Lancet Microbe. 2021;2(10):e508–e17.

[22.] Zheng H-Y, Xu M, Yang C-X, Tian R-R, Zhang M, Li J-J, et al. Longitudinal transcriptome analyses show robust T cell immunity during recovery from COVID-19. Signal Transduction and Targeted Therapy. 2020;5(1):294.

[23.] WHO R&D Blueprint novel Coronavirus COVID-19 Therapeutic Trial Synopsis. Geneva, Switzerland; 2020 18th February 2020.

[24.] R Development Core Team. R: A Language and Environment for Statistical Computing. 4.0.4 ed. Vienna, Austria 2020.

[25.] Zhang Y, Parmigiani G, Johnson WE. ComBat-seq: batch effect adjustment for RNA-seq count data. NAR Genomics and Bioinformatics. 2020;2(3).

[26.] Love MI, Huber W, Anders S. Moderated estimation of fold change and dispersion for RNA-seq data with DESeq2. Genome Biology. 2014;15(12):550.

[27.] Conesa A, Nueda MJ, Ferrer A, Talón M. maSigPro: a method to identify significantly differential expression profiles in time-course microarray experiments. Bioinformatics. 2006;22(9):1096–102.

[28.] Benjamini Y, Hochberg Y. Controlling the False Discovery Rate: A Practical and Powerful Approach to Multiple Testing. Journal of the Royal Statistical Society: Series B (Methodological). 1995;57(1):289–300.

[29.] Qin C, Zhou L, Hu Z, Zhang S, Yang S, Tao Y, et al. Dysregulation of Immune Response in Patients With Coronavirus 2019 (COVID-19) in Wuhan, China. Clinical Infectious Diseases. 2020;71(15):762–8.

[30.] Jackson H, Calle IR, Broderick C, Habgood-Coote D, d’Souza G, Nichols S, et al. Characterisation of the blood RNA host response underpinning severity in COVID-19 patients. medRxiv. 2021:2021.09.16.21263170.

[31.] Huang R, Meng T, Zha Q, Cheng K, Zhou X, Zheng J, et al. The predicting roles of carcinoembryonic antigen and its underlying mechanism in the progression of coronavirus disease 2019. Critical Care. 2021;25(1):234.

[32.] Zuo Y, Yalavarthi S, Shi H, Gockman K, Zuo M, Madison JA, et al. Neutrophil extracellular traps in COVID-19. JCI insight. 2020;5(11).

[33.] Barnes BJ, Adrover JM, Baxter-Stoltzfus A, Borczuk A, Cools-Lartigue J, Crawford JM, et al. Targeting potential drivers of COVID-19: Neutrophil extracellular traps. The Journal of experimental medicine. 2020;217(6).

[34.] Tang N, Li D, Wang X, Sun Z. Abnormal coagulation parameters are associated with poor prognosis in patients with novel coronavirus pneumonia. Journal of thrombosis and haemostasis : JTH. 2020;18(4):844–7.

[35.] Malas MB, Naazie IN, Elsayed N, Mathlouthi A, Marmor R, Clary B. Thromboembolism risk of COVID-19 is high and associated with a higher risk of mortality: A systematic review and meta-analysis. EClinicalMedicine. 2020;29:100639.

[36.] Hottz ED, Azevedo-Quintanilha IG, Palhinha L, Teixeira L, Barreto EA, Pão CRR, et al. Platelet activation and platelet-monocyte aggregate formation trigger tissue factor expression in patients with severe COVID-19. Blood. 2020;136(11):1330–41.

[37.] Shen S, Zhang J, Fang Y, Lu S, Wu J, Zheng X, et al. SARS-CoV-2 interacts with platelets and megakaryocytes via ACE2-independent mechanism. Journal of Hematology & Oncology. 2021;14(1):72.

[38.] Manne BK, Denorme F, Middleton EA, Portier I, Rowley JW, Stubben C, et al. Platelet gene expression and function in patients with COVID-19. Blood. 2020;136(11):1317–29.

[39.] Zaid Y, Puhm F, Allaeys I, Naya A, Oudghiri M, Khalki L, et al. Platelets Can Associate with SARS-Cov-2 RNA and Are Hyperactivated in COVID-19. Circulation research. 2020;127(11):1404–18.

[40.] Chen Y, Tong X, Li Y, Gu B, Yan J, Liu Y, et al. A comprehensive, longitudinal analysis of humoral responses specific to four recombinant antigens of SARS-CoV-2 in severe and non-severe COVID-19 patients. PLoS pathogens. 2020;16(9):e1008796.

[41.] Lucas C, Klein J, Sundaram ME, Liu F, Wong P, Silva J, et al. Delayed production of neutralizing antibodies correlates with fatal COVID-19. Nature Medicine. 2021;27(7):1178–86.

[42.] Zhang J-Y, Wang X-M, Xing X, Xu Z, Zhang C, Song J-W, et al. Single-cell landscape of immunological responses in patients with COVID-19. Nature Immunology. 2020;21(9):1107–18.

[43.] Chuprin A, Gal H, Biron-Shental T, Biran A, Amiel A, Rozenblatt S, et al. Cell fusion induced by ERVWE1 or measles virus causes cellular senescence. Genes & development. 2013;27(21):2356–66.

[44.] Martínez I, García-Carpizo V, Guijarro T, García-Gomez A, Navarro D, Aranda A, et al. Induction of DNA double-strand breaks and cellular senescence by human respiratory syncytial virus. Virulence. 2016;7(4):427–42.

[45.] Wiley CD, Liu S, Limbad C, Zawadzka AM, Beck J, Demaria M, et al. SILAC Analysis Reveals Increased Secretion of Hemostasis-Related Factors by Senescent Cells. Cell reports. 2019;28(13):3329–37.e5.

[46.] Coppé JP, Desprez PY, Krtolica A, Campisi J. The senescence-associated secretory phenotype: the dark side of tumor suppression. Annual review of pathology. 2010;5:99–118.

[47.] Lee S, Yu Y, Trimpert J, Benthani F, Mairhofer M, Richter-Pechanska P, et al. Virus-induced senescence is a driver and therapeutic target in COVID-19. Nature. 2021;599(7884):283–9.

[48.] Banerjee AK, Blanco MR, Bruce EA, Honson DD, Chen LM, Chow A, et al. SARS-CoV-2 Disrupts Splicing, Translation, and Protein Trafficking to Suppress Host Defenses. Cell. 2020;183(5):1325–39.e21.

[49.] Thoms M, Buschauer R, Ameismeier M, Koepke L, Denk T, Hirschenberger M, et al. Structural basis for translational shutdown and immune evasion by the Nsp1 protein of SARS-CoV-2. Science. 2020;369(6508):1249–55.

[50.] Au-Yeung BB, Shah NH, Shen L, Weiss A. ZAP-70 in Signaling, Biology, and Disease. Annual review of immunology. 2018;36:127–56.

[51.] Alon D, Paitan Y, Robinson E, Ganor N, Lipovetsky J, Yerushalmi R, et al. Downregulation of CD45 Signaling in COVID-19 Patients Is Reversed by C24D, a Novel CD45 Targeting Peptide. Frontiers in medicine. 2021;8:675963.

[52.] Jubel JM, Barbati ZR, Burger C, Wirtz DC, Schildberg FA. The Role of PD-1 in Acute and Chronic Infection. Frontiers in immunology. 2020;11.

[53.] Sturza A, Olariu S, Ionică M, Duicu OM, Văduva AO, Boia E, et al. Monoamine oxidase is a source of oxidative stress in obese patients with chronic inflammation (1). Canadian journal of physiology and pharmacology. 2019;97(9):844–9.

[54.] Nicotra A, Pierucci F, Parvez H, Senatori O. Monoamine oxidase expression during development and aging. Neurotoxicology. 2004;25(1-2):155–65.

[55.] Ionică LN, Gaiţă L, Bînă AM, Soșdean R, Lighezan R, Sima A, et al. Metformin alleviates monoamine oxidase-related vascular oxidative stress and endothelial dysfunction in rats with diet-induced obesity. Molecular and cellular biochemistry. 2021;476(11):4019–29.

[56.] Lighezan R, Sturza A, Duicu OM, Ceausu RA, Vaduva A, Gaspar M, et al. Monoamine oxidase inhibition improves vascular function in mammary arteries from nondiabetic and diabetic patients with coronary heart disease. Canadian journal of physiology and pharmacology. 2016;94(10):1040–7.

[57.] Sturza A, Leisegang MS, Babelova A, Schröder K, Benkhoff S, Loot AE, et al. Monoamine oxidases are mediators of endothelial dysfunction in the mouse aorta. Hypertension (Dallas, Tex : 1979). 2013;62(1):140–6.

[58.] Sturza A, Popoiu CM, Ionică M, Duicu OM, Olariu S, Muntean DM, et al. Monoamine Oxidase-Related Vascular Oxidative Stress in Diseases Associated with Inflammatory Burden. Oxidative medicine and cellular longevity. 2019;2019:8954201.

[59.] Cuperlovic-Culf M, Cunningham EL, Teimoorinia H, Surendra A, Pan X, Bennett SAL, et al. Metabolomics and computational analysis of the role of monoamine oxidase activity in delirium and SARS-COV-2 infection. Scientific Reports. 2021;11(1):10629.

[60.] Hok L, Rimac H, Mavri J, Vianello R. Relationship between COVID-19 infection and neurodegeneration: Computational insight into interactions between the SARS-CoV-2 spike protein and the monoamine oxidase enzymes. bioRxiv. 2021:2021.08.30.458208.

[61.] Zheng Z, Peng F, Xu B, Zhao J, Liu H, Peng J, et al. Risk factors of critical & mortal COVID-19 cases: A systematic literature review and meta-analysis. The Journal of infection. 2020;81(2):e16–e25.

[62.] Wolff D, Nee S, Hickey NS, Marschollek M. Risk factors for Covid-19 severity and fatality: a structured literature review. Infection. 2021;49(1):15–28.

